# Role of FYVE and Coiled-Coil Domain Autophagy Adaptor 1 in severity of COVID-19 infection

**DOI:** 10.1101/2021.01.22.21250070

**Authors:** Sandra P. Smieszek, Bartlomiej Przychodzen, Christos Polymeropoulos, Vasilios Polymeropoulos, Mihael H. Polymeropoulos

## Abstract

Coronaviruses remodel intracellular membranes to form specialized viral replication compartments, such as double-membrane vesicles where viral RNA genome replication takes place. Understanding the factors affecting host response is instrumental to design of therapeutics to prevent or ameliorate the course of infection.

As part of explorative tests in hospitalized patients with confirmed COVID-19 infection participating in ODYSSEY trial, we obtained samples for whole genome sequencing analysis as well as for viral genome sequencing. Based on our data, we confirm one of the strongest severity susceptibility locus thus far reported in association with severe COVID-19: 3p21.31 locus with lead variant rs73064425. We further examine the associated region. Interestingly based on LD analysis we report 3 coding mutations within one gene in the region of FYVE and Coiled-Coil Domain Autophagy Adaptor 1 (*FYCO1*). We specifically focus on the role of *FYCO1* modifiers and gain-of-function variants. We report the associations between the region and clinical characteristics in this severe set of COVID-19 patients.

We next analyzed expression profiles of *FYCO1* across all 466 compounds tested. We selected only those results that showed a significant reduction of expression of *FYCO1*. The most significant candidate was indomethacin – an anti-inflammatory that could potentially downregulate *FYCO1.* We hypothesize that via its direct effects on efficiency of viral egress, it may serve as a potent therapeutic decreasing the replication and infectivity of the virus. Clinical studies will be needed to examine the therapeutic utility of indomethacin and other compounds downregulating *FYCO1* in COVID-19 infection and other strains of betacoronaviruses.

## Introduction

The emergence of highly pathogenic zoonotic viruses calls for further research in order to understand, treat and ideally prevent such infections ultimately circumventing epidemics. Currently accurate prognoses are confounded by our inability to predict disease severity and virulence across space and time. Virus-human interactions have driven around 30% of the human genome evolution since divergence from chimpanzee. Different variants, genes affect severity to a range of infections, for example: *IFITM3, IRF*7 in influenza, *SFPA/D* and *VTR* in RSV, *CCR5* in HIV, *IFNG* in HTVL-1 and *TNF* in HPV to name a few. The complex interplay between environmental, viral, and host genetic factors drives differences in individual disease progression, severity, and outcome results. Understanding resilience and susceptibility factors should enable one to design prophylactics and therapeutics.

SARS-CoV-2 is a novel coronavirus that has led to a worldwide pandemic^1^. There is a high mortality rate associated with the virus and despite measures implemented to contain the spread of the virus, over 2.1 million deathswere reported worlwide (1/26/21). The ACE2 binding affinity with RBD on the Spike protein of SARS-CoV-2 is 10-20x higher than that of the SARS-CoV. The clinical spectrum of this disease is heterogeneous, and infected individuals viral RNA shedding pattern differ across individuals at baseline. Similarly to other positive-sense RNA viruses, coronaviruses remodel intracellular membranes to form specialized viral replication compartments. Depending on the time course of the infection, the structure, composition, and formation of virus replication organelles appear to be varied and dynamic. Understanding the mode of action of this virus in the context of its interaction with the host genome is fundamental to the design of optimal therapeutic strategies.

In the early months of the epidemic it became apparent that advanced age and comorbidities are associated with higher risk for severe infection yet none of these fully explain the heterogeneous course of the infection amongst individuals^2^. To that end several sequencing projects have been commenced since the beginning of the epidemic. Initial results from COVID-19 initiative suggest a locus on chromosome 3p21.31, the peak association with severity of the disease, signal that has been observed by the Severe Covid-19 GWAS Group and further replicated by other host genome sequencing consortia^345^ and the UKbiobank^6^. The genetic variants on chromosome 3 that are most associated with severe COVID-19 are all in high linkage disequilibrium (LD). Furthermore recent phylogenetic analysis showed that the risk haplotype thus entered the modern human population from Neanderthals (Vindija 33.19 Neanderthal)^7^. Specifically, we focused on this region is because the 3p21.31 regional severity association was the strongest one replicated by multiple consortia including the COVID-19 initiative and latest Nature report by Casteneira et al., (rs73064425, OR=2.14, discovery p=4.77 × 10-30) (top variants can be accessed in the supplementary material)

As part of explorative tests in hospitalized patients with confirmed SARS-CoV-2 positive infection participating in ODYSSEY trial, we obtained blood samples for whole genome sequencing analysis as well as nasopharyngeal swabs for further viral genome sequencing. We have conducted whole genome sequencing on first cohort participants as well as deep clinical phenotyping. Based on our data, we confirm one of the strongest susceptibility locus reported and now replicated with lead variant being rs73064425. We further examine the associated region, with specific focus on the role of *FYCO1* gene. We further discuss the associations between the region and clinical characteristics in this severe set of COVID-19 patients.

## Results

### Regional association

We replicate the strongest susceptibility locus reported and now replicated is within, *3p21.31 locus,* rs73064425 Leucine Zipper Transcription Factor Like 1 (LZTFL1) reported by Casteneira et al., as the strongest association with severity (rs73064425, OR=2.14, discovery p=4.77 × 10-30). We replicate this association in our cohort of hospitalized patients (OR 3.2, CI 1.8728 to 5.4637, p-value<0.0001). The variant has a global gnomAD MAF of 0.05. We report an increased MAF 0.19 in ODYSSEY, and consistently with gnomAD, we report a MAF of 0.05 in Vanda 2000 controls. The highest allelic frequency for this variant globally is present in Ashkenazi Jewish (MAF 0.12) and the lowest frequency is present amongst East and South Asian populations (MAF 0.0006). The GWAS signal covers a cluster of six genes (*SLC6A20, LZTFL1, CCR9, FYCO1, CXCR6*, and *XCR1*), several of which with functions that could be potentially relevant to Covid-19. The variant is a strong eQTL for CXCR6 and SLC6A20 in GTEX^8^. The locus itself is within a highly regulatory region, as an enhancer for Primary B cells from peripheral blood and modifier H3K4me1 in accordance with Haploreg. Furthermore the locus is associated with monocyte percentage (UK Biobank field: 30190-0.0) in Gene Atlas.

Next we reconstructed LD to test if the signal (non-coding, top SNP by p-value) from other large studies can be reproduced and extended to nearby genes, Where are the “limits” of the signal on Odyssey data, Are there any coding variants .Interestingly there are 3 coding mutations within one gene in the region (*FYCO1*) causing 2 amino acid substitutions (Table1; two mutations within same codon, 1001, resulting in one missense mutation, third in strong LD). All 3 coding variants are in strong LD with top SNPs published previously within the region and seem to be the strongest signal for our Odyssey samples. Having ∼2000 control genomes we can see that all 3 coding variants form a strong LD. Quintessentially the risk is conveyed by the extent of existing LD structure in that region as shown in Supplementary Table1. Specifically, rs33910087 is an eQTL for CXCR6 and based on GeneAtlas, the genotyped variant is highly significant modifier of monocyte percentage (p=3.8479e-46).

**Table 1.** Top coding variants in association with severity of COVID19 infection.

| CHR | BP | SNP | A1 | MAF.Covid19 | MAF.Controls | Pval | OR | Ref | Alt | Gene | Function | AAChange | RSID |
| --- | --- | --- | --- | --- | --- | --- | --- | --- | --- | --- | --- | --- | --- |
| 3 | 46007825 | 3:46007825[b37]C,T | C | 22.3% | 8.3% | 7.01E-06 | 3.168 | T | C | FYCO1 | nonsynonymous | c.A3001G:p.N1001D | rs13059238 |
| 3 | 46007823 | 3:46007823[b37]G,T | T | 21.4% | 7.8% | 7.84E-06 | 3.22 | G | T | FYCO1 | nonsynonymous | c.C3003A:p.N1001K | rs13079478 |
| 3 | 46009487 | 3:46009487[b37]A,G | A | 21.4% | 8.1% | 1.34E-05 | 3.104 | G | A | FYCO1 | nonsynonymous | c.C1339T:p.R447C | rs33910087 |
| 3 | 45901089 | 3:45901089[b37]C,T | T | 17.0% | 5.3% | 1.19E-05 | 3.628 | C | T | LZTFL1 | intronic | . | rs73064425 |

The association of the locus with severity of infection in interaction with SARS-CoV-2 remains to be examined. Functionally *FYCO1* encodes a protein involved in vesicle transport and autophagy. It has been suggested as a key mediator linking ER-derived double membrane vesicles, the primary replication site for coronaviruses, with the microtubule network^9^. *FYCO1* through its LC3-interacting region (LIR) motif was shown to be important for the fusion of autophagosomes with lysosomes. *FYCO1* dimerizes via the CC region, interacts with PI3P via its FYVE domain, and forms a complex with Rab7 via a part of the CC region located in front of the FYVE domain^9^. Specifically, *FYCO1* was shown to act as a Rab7 effector that binds to LC3 and PI3P to mediate microtubule plus end-directed vesicle transport^10^. The depletion of Rab7 inhibits maturation of late endosomes/MVBs and leads to reduced lysosome numbers in cells^11^. *FYCO1* also mediates clearance of α□synuclein aggregates through a Rab7□dependent mechanism, overexpression reducing number of cell with α□synuclein aggregates^12^. In another recent report authors perform a genome-scale CRISPR loss-of-function screen in human alveolar basal epithelial carcinoma cells to identify genes whose loss enabled resistance to SARS-CoV-2 viral infection^13^. Loss of RAB7A reduces viral entry/egress by sequestering the ACE2 receptor inside cells^13^. Furthermore, depletion of *FYCO1* or antibodies against the N-terminus of LC3 blocks the subcellular redistribution of autophagosomes^14^. Indeed, rare *FYCO1* variants some of which contain missense mutations in the LIR domain have recently been associated with inclusion body myositis a disease characterized by impaired autophagic degradation.

We stipulate gain of function variants in *FYCO1* confer higher risk of severe course of COVID-19 induced infection and likely other betacoronaviruses. Temporarily downregulating *FYCO1* seems to be protective, hence the locus is likely crucial to designing therapeutic strategies for this and other strain of betacoronaviruses.

### FYCO1 and drug-gene expression

We have conducted a high throughput drug screen gene expression analysis to identify compounds that would downregulate the expression of *FYCO1*. To discover potential, pharmaceutical agents capable of affecting transcriptional expression levels of *FYCO1* implicated in SARS-CoV and SARS-CoV2 patho-physiology, we have screened 466 compounds belonging to 14 different therapeutic classes. Screening was conducted using human retinal pigment epithelia cell line (ARPE-19) and gene expression changes were collected across 12,490 genes. We analyzed the expression profiles of *FYCO1* across all 466 compounds tested. In order to find positive hits we selected only those results that showed a reduction of expression of *FYCO1* (1.5 -fold difference). The top significant candidate indomethacin is (displayed in **Table 2**) a drug that has both anti-inflammatory and antiviral actions. Indomethacin was previously shown to have potent direct antiviral activity against the coronaviruses SARS-CoV and CCoV. Its potent antiviral activity (>1,000-fold reduction in virus yield) was previously confirmed in vivo in CCoV-infected dogs (Amici). These screens warrant further confirmatory tests nevertheless the strategy to weaken viral egress could be potentially useful for strains beyond SARS-CoV-2

**Table 2.**
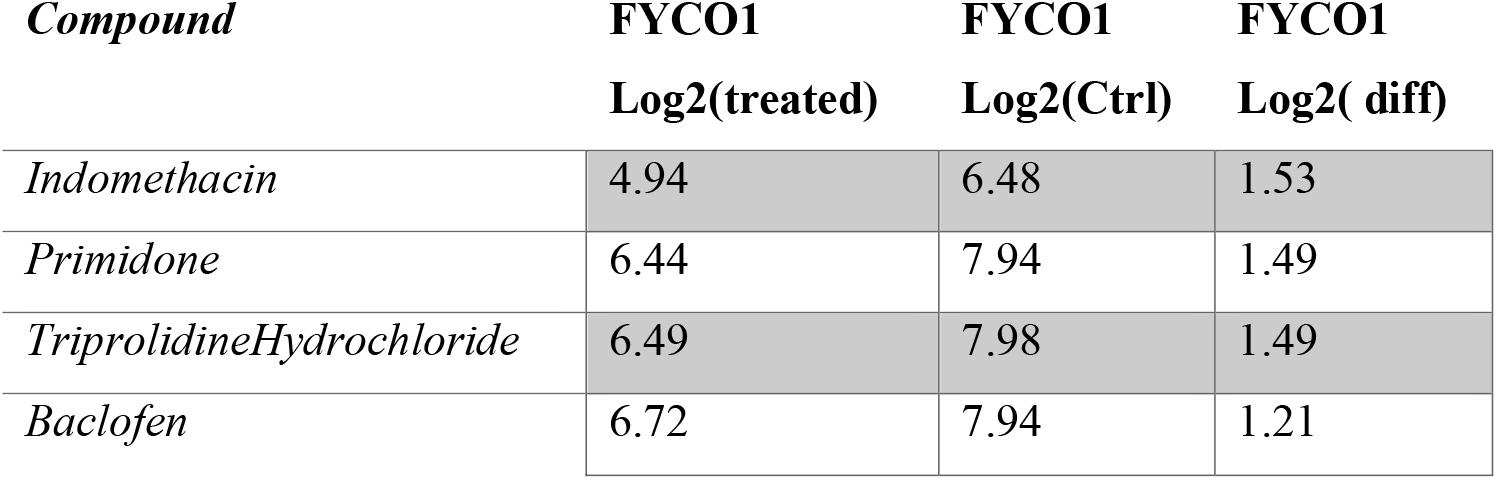
Top significant candidates.

### Clinical Characteristics of carriers

Both heterozygotes and homozygotes were severe hospitalized patients treated for COVID-19. The signal is one of severity however we wanted to check if the variant carriers would have any particular clinical characteristics. To that end we tested systemic cytokine panel in addition to standard lab tests. Clinical characteristics across genotypes are displayed in **Supplementary Material**. Noticeable is the linear distribution with higher sodium and chloride seen in homozygous individuals. All study patients met inclusion criteria included: age 18-90; confirmed laboratory COVID-19 infection; confirmed pneumonia by chest radiograph or computed tomography; fever defined as temperature ≥ 36.6 °C armpit, ≥ 37.2 °C oral, or ≥ 37.8 °C rectal since admission or the use of antipyretics; PaO2 / FiO2 ≤ 300; 6. in-patient hospitalization. We furthermore sequenced the viral genome sequence data in conjunction with host sequencing. We report coding mutations in SARS-CoV-2 based CoV-GLUE annotations^15^. All known variants reported in CoV-Glue^15^ (until 11/23/2020) variants were annotated. As the viral strains evolved, the 614th aa position of the S protein, aspartate became replaced by glycine ultimately becoming the ubiquitous strain, hence making open conformations more likely. All patients are carriers of the S:D614G variant -variant in the spike protein D614G that rapidly became dominant strain according to latest GISAID data^16,17^.

## Discussion

Coronaviruses remodel intracellular membranes to form specialized viral replication compartments, such as double-membrane vesicles where viral RNA genome replication takes place. Understanding the formation and operation of these vesicles is instrumental to optimal design of therapeutics to prevent or ameliorate the course of infection. Here via findings from whole genome sequencing and association with severity of COVID-19, we were led to a locus that may in fact be functionally relevant in predisposing individuals to severe course of infection via its direct effects on efficiency of viral egress.

*FYCO1* has a direct link to formation of vesicles and autophagy. Pathogenic mutations in FYCO1 can affect intracellular transport of autophagocytic vesicles. Depletion of *FYCO1* has been shown to block distribution of autophagosomes. *FYCO1* also mediates clearance of α□synuclein aggregates through a Rab7□dependent mechanism, overexpression reducing number of cell with α□synuclein aggregates. Recently using single cell RNAseq authors identified a group of genes (ATP6AP1, ATP6V1A, NPC1, RAB7A, CCDC22, and PIK3C3) whose knockout induced shared transcriptional changes in cholesterol biosynthesis pathway^13^.

They showed how the actual perturbation of the cholesterol biosynthesis pathway reduced viral infection. That furthermore reveals the links via RAB7. We hypothesize fewer pLOFs in NPC1 to be present in cohorts of severe hospitalized patients with COVID-19. Niemann-Pick disease type C1 lipid storage disorder offers resistance to Ebola in cell line experiments and in homozygous recessive mice ((Npc1-/-))^18^,^19^. Furthermore, bat species show selective sensitivity to Ebola versus Marburg viruses^20^. Our hypothesis is that people with certain lysosomal storage diseases may be resistant to one of these viruses. Along these lines there is suggestive evidence for this to be exactly the case. This is in line with a report by Sturley et al., authors propose that 50 loci that in the homozygous state cause a lysosomal storage disorder, may account for divergent clinical outcomes in COVID-19.

Blocking or perturbing viral egress whether via Rab7, FYCO1 or other components may be a viable strategy to stop the proliferation of the virus reducing viral load. In the case of betacoronaviruses unconventional egress can be blocked by the Rab7 GTPase competitive inhibitor CID1067700^11^ -compound potently decreasing viral egress in a dose-dependent manner. Here we focused on downregulation of FYCO1: based on cell line expression with resulting potential candidates including indomethacin -a drug that has both anti-inflammatory and antiviral actions. Slowing viral spread by targeting regulators of lysosomal trafficking, by focusing on egress and double membrane vesicles may offer a therapeutics strategy for novel strain of betacoronaviruses.

The GWAS locus has led for further understanding of not only the more efficient egress leading to successful propagation of the viral particles but also to potential protective mechanisms and greater understanding of the heterogeneous clinical outcomes. We hence stipulate gain of function variants in *FYCO1* confer higher risk of infection with SARS and other betacoronaviruses. Downregulating or temporarily limiting, *FYCO1* activity may be protective. This susceptibility locus in fact highlights the potentially druggable cellular aspects of the host.

## Methods

### Clinical Trial Information

ODYSSEY is a double-blinded Phase 3 study with a planned randomization of a total of 300 hospitalized severely ill COVID-19 patients to receive either tradipitant 85 mg bid or placebo for a total of up to 14 days or discharge (CONSORT Flowchart in S. File 1). The randomization is stratified by site with a block size of four. Inclusion criteria for the study comprised of: 1. Adults aged 18-90; 2. confirmed laboratory COVID-19 infection; 3. confirmed pneumonia by chest radiograph or computed tomography; 4. fever defined as temperature ≥ 36.6 °C armpit, ≥ 37.2 °C oral, or ≥ 37.8 °C rectal since admission or the use of antipyretics; 5. PaO2 / FiO2 ≤ 300; 6. in-patient hospitalization. Patients were to be followed for up to 28 days to record clinical outcomes. Patients’ clinical progress was recorded on a 7 point clinical status ordinal scale defined as follows: 1-Death; 2-Hospitalized on mechanical ventilation or ECMO; 3-Hospitalized on non-invasive ventilation or high-flow oxygen supplementation; 4-Hospitalized requiring supplemental oxygen; 5-Hospitalized not requiring supplemental oxygen, requiring continued medical care; 6-Hospitalized not requiring supplemental oxygen, not requiring continued medical care; 7-Not hospitalized.

### Human whole genome sequencing

#### Vanda Pharmaceuticals Inc. COVID-19 Cohort

WGS cohort consisted of 80 COVID-19 hospitalized patient samples and 1876 WGS controls. DNA was extracted from 200 µl of whole blood using QIAamp blood mini kit and eluted into 100 µl volume in order to obtain 500 ng of DNA. Libraries were prepared by random fragmentation of DNA followed by adapter ligation using Illumina TruSeq adapters. Paired-end libraries 2×150 bp were sequenced on an Illumina NovaSeq6000 S4 with a target of 90Gb raw read depth per sample. The cohort was multi-ethnic and the age ranged from 35-87 for severe hospitalized COVID-19 cases (68% male, 32% female). Specifically the PC defined ancestry was as follows (African 9.8%, European 46.3%, Asian 4.9%, Hispanic 22% and other 17.1%). Furthermore, 16% of the cohort were patients hospitalized on mechanical ventilation or ECMO, 13% deceased.

Both cases and controls were processed with the same bioinformatic pipeline for variant calling. Paired-end 150□Jbp reads were aligned to the GRCh37 human reference (BWA-MEM v0.7.8) followed by Picard (MarkDuplicates) and processed with GATK best-practices workflow (GATK v3.5.0). The mean coverage was 35.8x. All high quality variants obtained from GATK were annotated. Annotations include variant effect predictions using VEP; allele frequencies from Gnomad;dbSNP 150 rsIDs; conservation scores from PhyloP, GERP, PhastCons; damaging effect predictions from Polyphen 2, SIFT and clinically relevant information from ClinVar. We removed samples with a discordance between self-declared and sequence-derived gender, we used KING for detection of related individuals and performed PCA for ancestry adjustment. The lead variant was in addition verified with genotyping (Supplementary Material). Logistic regression was conducted in PLINK, MATLAB. The analysis was corrected for PC 1-15, age and sex.

#### Viral sequencing and genome assembly

Sequencing was attempted on all samples with a positive RT-PCR assay result that had Ct <32 using either a metagenomic approach described previously^21^, via IDT probe-capture^22^, or by amplicon sequencing with SWIFT library preparation. Libraries were sequenced on Illumina MiSeq or NextSeq instruments using 1×150 or 1×75 runs respectively. Consensus sequences were assembled using a custom bioinformatics pipeline [https://github.com/proychou/hCoV19] adapted for SARS-CoV-2 from previous work^22^,^23^. Briefly, raw reads were trimmed to remove adapters and low quality regions using BBDuk and a k-mer based filter was used to pull out reads matching the reference sequence NC_045512. Filtered reads were *de novo* assembled using SPAdes^24^ and contigs were ordered against the reference using BWA-MEM. Gaps were filled by remapping reads against the assembled scaffold and a consensus sequence was called from this alignment using a custom script in R/Bioconductor.

#### Cell culture and drug treatment

Drugs screening was carried out, the same one as applied in our previous study^25^. The retinal pigment epithelia cell line, ARPE-19/HPV-16, was chosen to establish a database of drug profiles because of its non-cancerous, human origin, with a normal karyotype. It can also be easily grown as monolayer in 96-well plates. Compounds were obtained from Sigma (St. Louis, MO) or Vanda Pharmaceuticals (Washington, DC). Cells were aliquoted on 96-well plates (∼2×10e5 cells/well) and incubated for 24 h prior to providing fresh media with a drug, or the drug vehicle (water, dimethyl sulfoxide, ethanol, methanol, or phosphate-buffered saline solution). Drugs were diluted 1000 fold in buffered in Dulbecco’s Modified Eagle Medium: Nutrient Mixture F-12 (D-MEM/F-12) culture medium (Invitrogen, Carlsbad, CA) containing nonessential amino acids and 110 mg/L sodium pyruvate. In these conditions, no significant changes of pH were expected, which was confirmed by the monitoring of the pH indicator present in the medium. A final 10 μM drug concentration was chosen because it is believed to fit in the range of physiological conditions^25^. Microscopic inspection of each well was conducted at the end of the treatment to discard any samples where cells had morphological changes consistent with apoptosis. We also verified that the drug had not precipitated in the culture medium.

#### Gene expression

Cells were harvested 24 h after treatment and RNA was extracted using the RNeasy 96 protocol (Qiagen, Valencia, CA). Gene expression for 22,238 probe sets of 12,490 genes was generated with U133A2.0 microarrays following the manufacturer’s instructions (Affymetrix, Santa Clara, CA). Drugs were profiled in duplicate or triplicate, with multiple vehicle controls on each plate. A total of 708 microarrays were analyzed including 74 for the 18 antipsychotics, 499 for the other 448 compounds, and 135 for vehicle controls. The raw scan data were first converted to average difference values using MAS 5.0 (Affymetrix). The average difference values of both treatment and control data were set to a minimum of 50 or lower. For each treatment category, all probe sets were then ranked based on their amplitude, or level of expression relative to the vehicle control (or the average of controls was selected when more than one was used). Amplitude was defined as the ratio of expression (t−v) / [(t+v) / 2] where t corresponds to treatment instance and v to vehicle instance. Each drug group profile was created using our novel Weighted Influence Model, Rank of Ranks (WIMRR) method which underscores the rank of each probe set across the entire gene expression profile rather than the specific change in expression level. WIMRR takes the average rank of each probe set across all of the members of the group and then reranks the probe sets from smallest average rank to largest average rank. A gene-set enrichment metric based on the Kolmogorov– Smirnov (KS) statistic. Specifically, for a given set of probes, the KS score gives a measure of how up (positive) or down (negative) the set of probes occurs within the profile of another treatment instance.

## Data Availability

available upon request

## Declarations

### Funding

Vanda Pharmaceuticals Inc.

### Competing Interests

SPS,BPP, CP. VP MHP are employees of Vanda Pharmaceuticals.

### Ethical Approval

Reviewed and approved by Advarra IRB; Pro00043096

---

**Figure 1.**
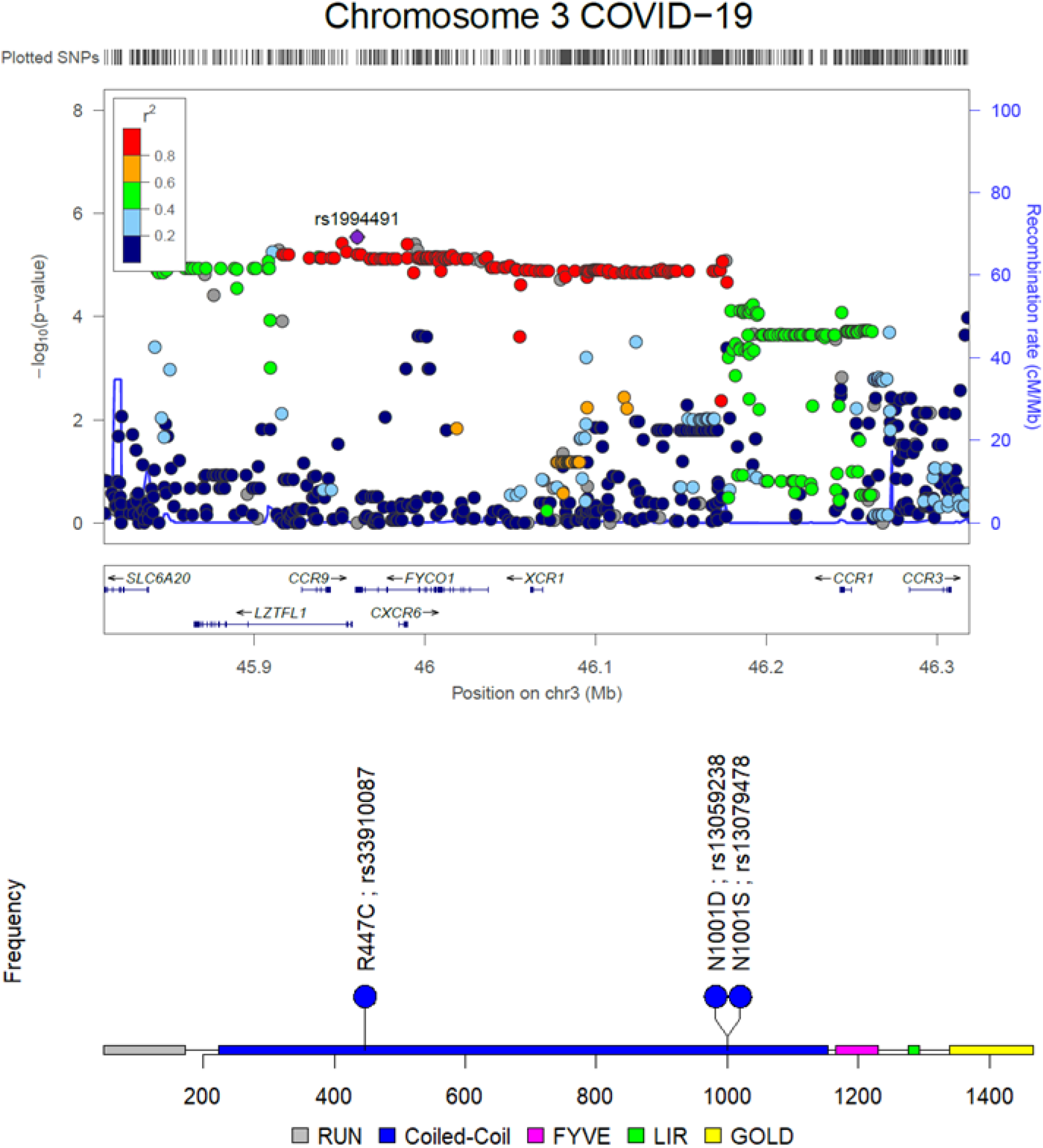
Figure 1A depicts the coding variants detected in the region association with severity of COVID-19 infection in FYCO1 gene. Figure 1B is a locus zoom of the entire significant chromosome 3 region associated with severe COVID-19 based on a whole genome sequencing analysis.

## Notes

### Competing Interest Statement

All work has been sponsored by Vanda Pharmaceuticals Inc.
All authors are employees of Vanda Pharmaceuticals Inc.

### Clinical Trial

NA

### Funding Statement

No funding

### Summary of Updates

minor corrections and Table 1 included,

