## Supplementary Material for "Role of FYVE and Coiled-Coil Domain Autophagy Adaptor 1 in severity of COVID-19 infection"

**Role of FYVE and Coiled-Coil Domain Autophagy Adaptor 1 in severity of COVID-19 infection: from GWAS hit to therapeutic hypothesis**

### LD association of top3 coding SNPs

| CHR_A | BP_A | SNP_A | CHR_B | BP_B | SNP_B | R2 |
|---|---|---|---|---|---|---|
| 3 | 46007823 | rs13079478 | 3 | 46007825 | rs13059238 | 0.943138 |
| 3 | 46007823 | rs13079478 | 3 | 46009487 | rs33910087 | 0.970857 |
| 3 | 46007825 | rs13059238 | 3 | 46009487 | rs33910087 | 0.971407 |

### COVID-19 severity GWAS
### Erola Pairo-Castineira et al., 2020

**Table 1 | Lead variants from independent genome-wide significant regions**

| SNP | chr:pos(b37) | Risk | Alt | $RAF_{gcc}$ | $RAF_{ukb}$ | OR | CI | $P_{gcc.ukb}$ | $P_{gcc.gs}$ | $P_{gcc.100k}$ | Locus |
|---|---|---|---|---|---|---|---|---|---|---|---|
| rs73064425 | 3:45901089 | T | C | 0.15 | 0.07 | 2.1 | 1.88-2.45 | $4.8 \times 10^{-30}$ | $2.9 \times 10^{-27}$ | $3.6 \times 10^{-32}$ | *LZTFL1* |
| rs9380142 | 6:29798794 | A | G | 0.74 | 0.69 | 1.3 | 1.18-1.43 | $3.2 \times 10^{-8}$ | 0.00091 | $1.8 \times 10^{-8}$ | *HLA-G* |
| rs143334143 | 6:31121426 | A | G | 0.12 | 0.07 | 1.9 | 1.61-2.13 | $8.8 \times 10^{-18}$ | $2.6 \times 10^{-24}$ | $5.8 \times 10^{-18}$ | *CCHCR1* |
| rs3131294 | 6:32180146 | G | A | 0.9 | 0.86 | 1.5 | 1.28-1.66 | $2.8 \times 10^{-8}$ | $1.3 \times 10^{-10}$ | $2.3 \times 10^{-8}$ | *NOTCH4* |
| rs10735079 | 12:113380008 | A | G | 0.68 | 0.63 | 1.3 | 1.18-1.42 | $1.6 \times 10^{-8}$ | $2.8 \times 10^{-9}$ | $4.7 \times 10^{-6}$ | *OAS1/3* |
| rs2109069 | 19:4719443 | A | G | 0.38 | 0.32 | 1.4 | 1.25-1.48 | $4 \times 10^{-12}$ | $4.5 \times 10^{-7}$ | $2.4 \times 10^{-8}$ | *DPP9* |
| rs74956615 | 19:10427721 | A | T | 0.079 | 0.05 | 1.6 | 1.35-1.87 | $2.3 \times 10^{-8}$ | $2.2 \times 10^{-13}$ | $3.9 \times 10^{-6}$ | *TYK2* |
| rs2236757 | 21:34624917 | A | G | 0.34 | 0.28 | 1.3 | 1.17-1.41 | $5 \times 10^{-8}$ | $8.9 \times 10^{-5}$ | $8.3 \times 10^{-7}$ | *IFNAR2* |

chr:pos - chromosome and position of the top SNP (build 37); Risk – risk allele; Alt - other allele; RAF - risk allele frequency; OR - effect size (odds ratio) of the risk allele in the GenOMICC EUR analysis; CI - 95% confidence interval for the odds ratio in the GenOMICC EUR cohort; P - p-value, Locus – gene nearest to the top SNP. Subscript identifiers indicate the cohorts used for cases: gcc - GenOMICC EUR; and controls: ukb - UK Biobank; gs - Generation Scotland; 100k - 100,000 genomes.

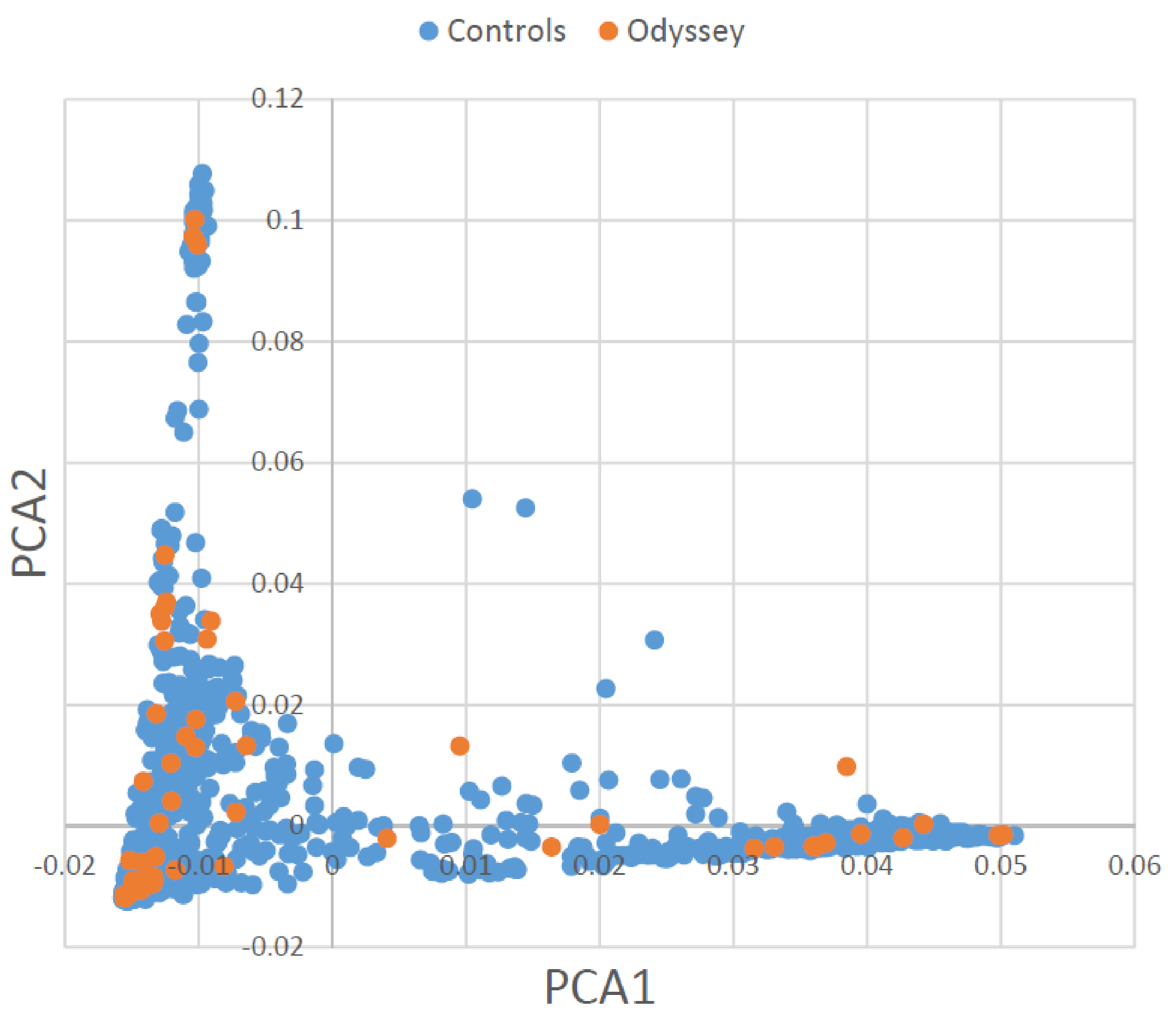

Controls
Odyssey
PCA2
PCA1
0.12
0.1
0.08
0.06
0.04
0.02
0
-0.02
-0.02
-0.01
0
0.01
0.02
0.03
0.04
0.05
0.06

### QQ plot

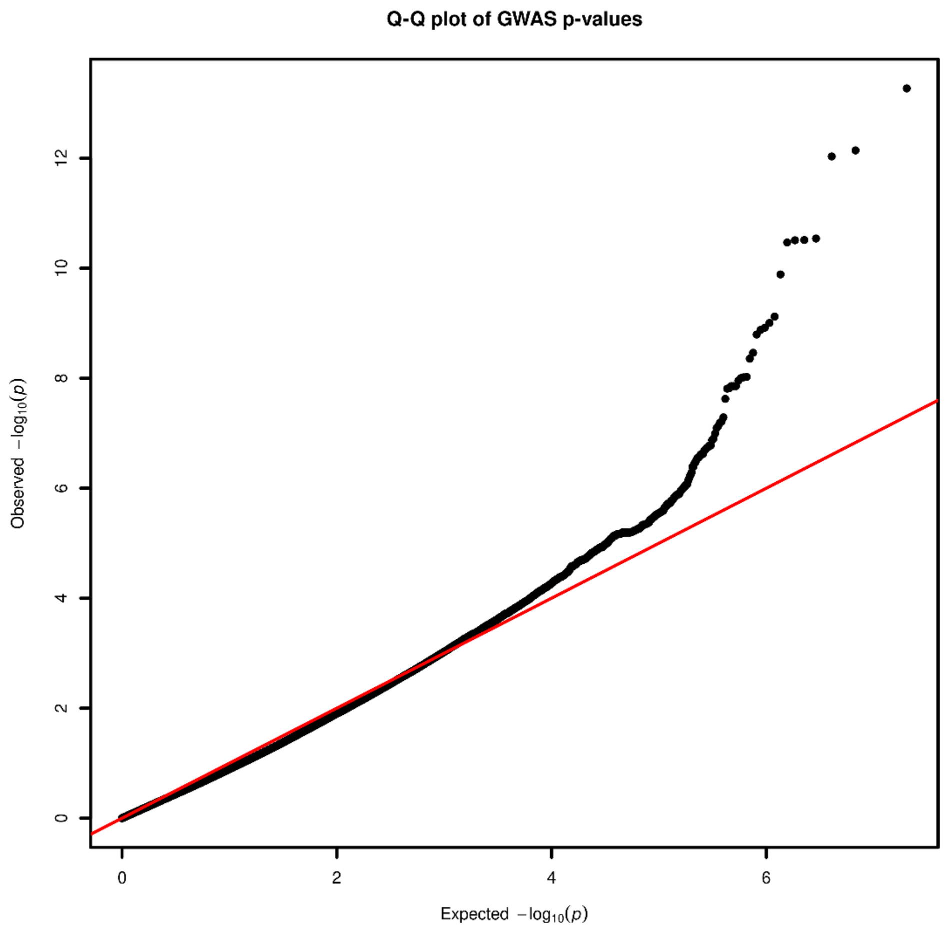
Q-Q plot of GWAS p-values
Observed $-\log_{10}(p)$
0
2
4
6
8
10
12
Expected $-\log_{10}(p)$
0
2
4
6

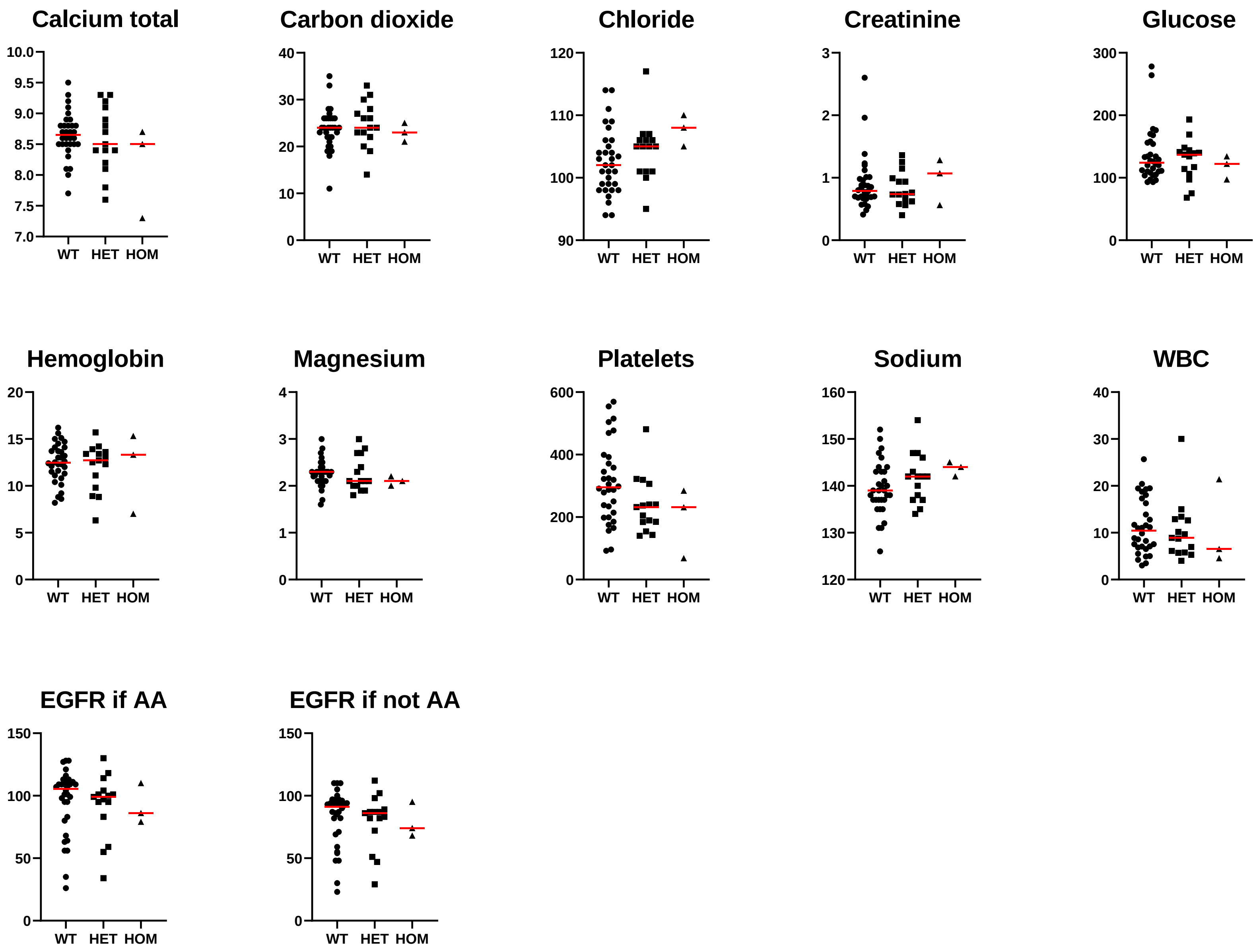
Calcium total
10.0
9.5
9.0
8.5
8.0
7.5
7.0
WT HET HOM
Carbon dioxide
40
30
20
10
0
WT HET HOM
Chloride
120
110
100
90
WT HET HOM
Creatinine
3
2
1
0
WT HET HOM
Glucose
300
200
100
0
WT HET HOM
Hemoglobin
20
15
10
5
0
WT HET HOM
Magnesium
4
3
2
1
0
WT HET HOM
Platelets
600
400
200
0
WT HET HOM
Sodium
160
150
140
130
120
WT HET HOM
WBC
40
30
20
10
0
WT HET HOM
EGFR if AA
150
100
50
0
WT HET HOM
EGFR if not AA
150
100
50
0
WT HET HOM

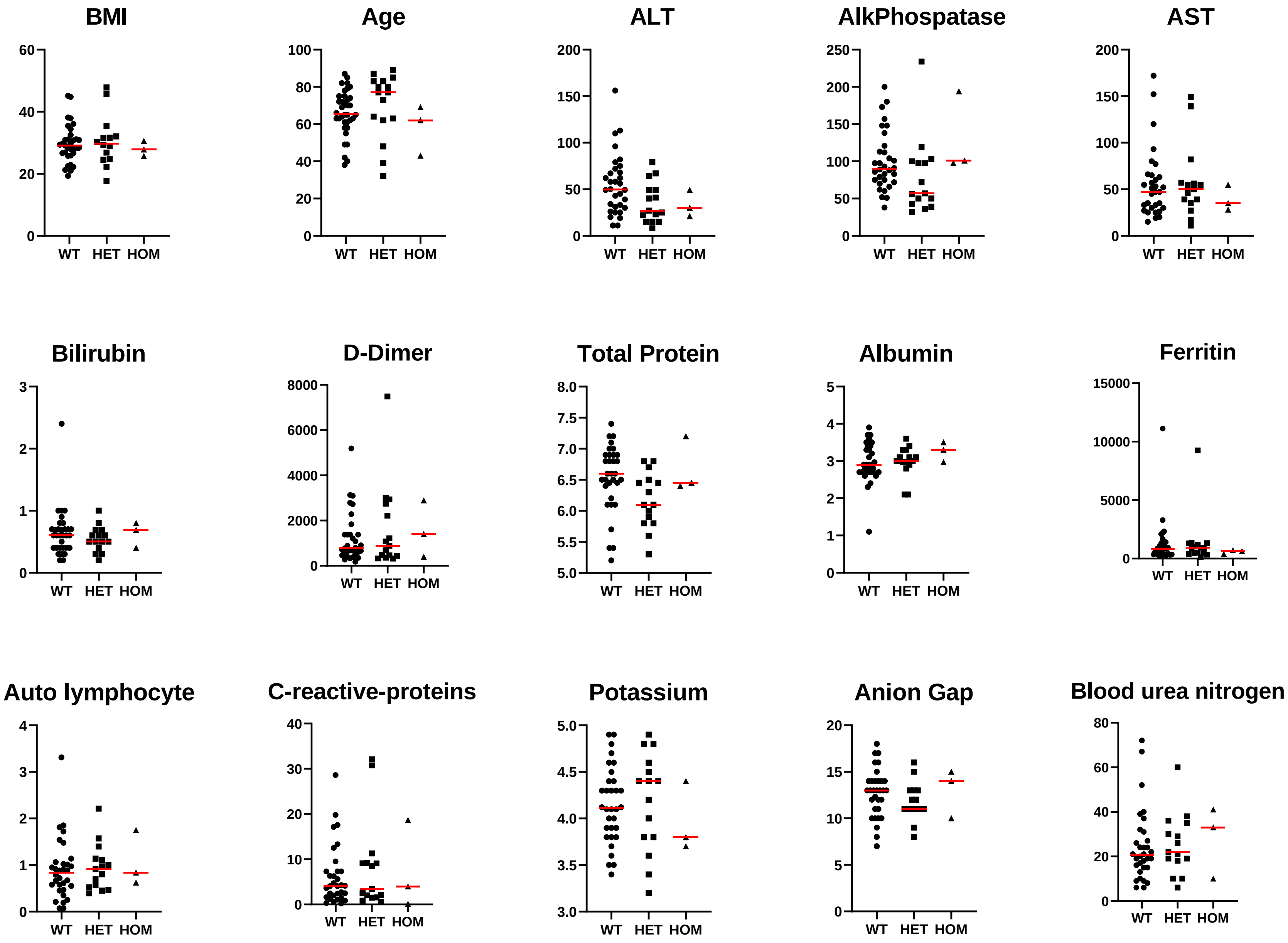
BMI
Age
ALT
AlkPhospatase
AST
Bilirubin
D-Dimer
Total Protein
Albumin
Ferritin
Auto lymphocyte
C-reactive-proteins
Potassium
Anion Gap
Blood urea nitrogen
WT
HET
HOM

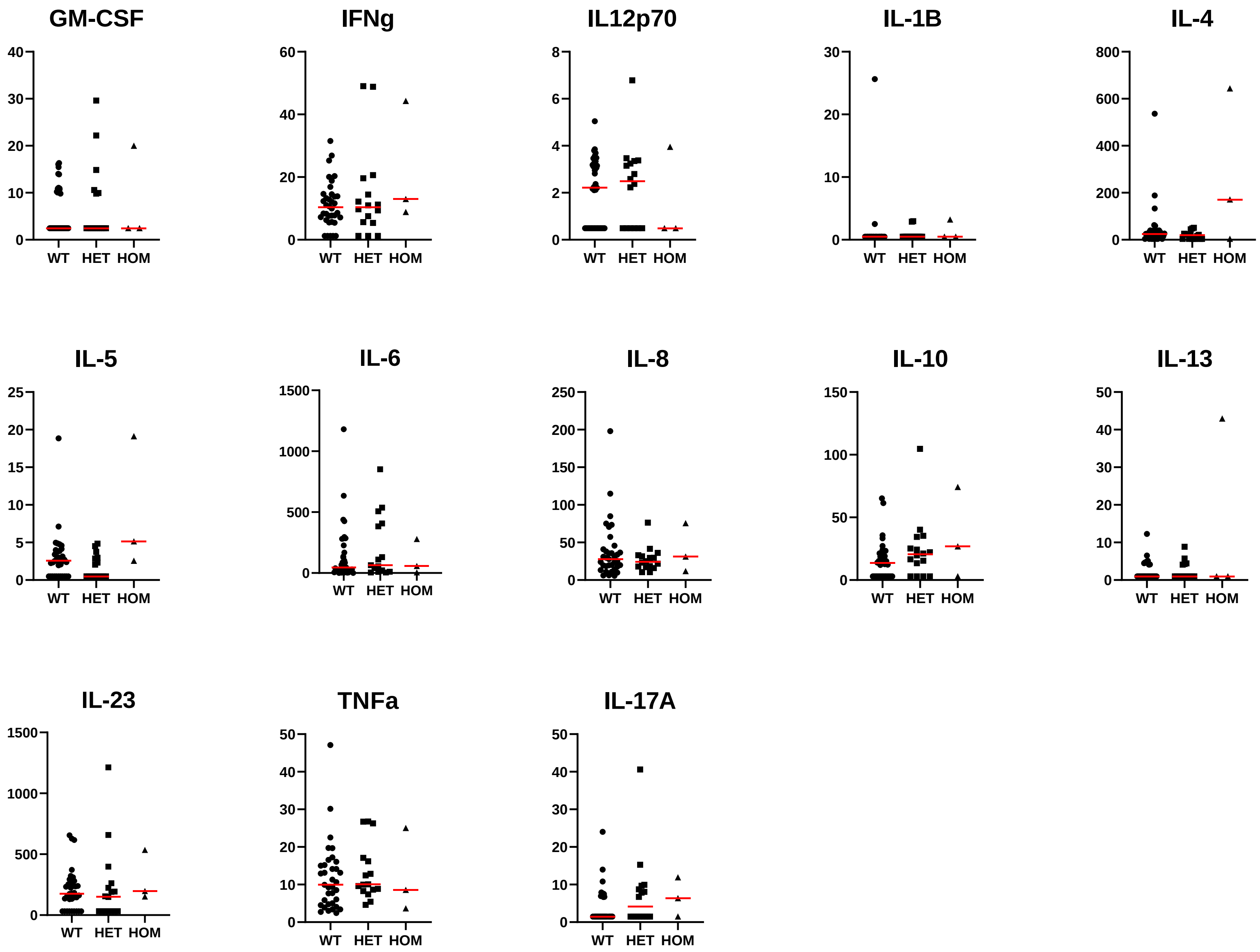
GM-CSF
IFNg
IL12p70
IL-1B
IL-4
IL-5
IL-6
IL-8
IL-10
IL-13
IL-23
TNFa
IL-17A
WT HET HOM

#### African

| RS Number | Position (GRCh37) | Allele Frequencies | Haplotypes | |
|---|---|---|---|---|
| rs73064425 | chr3:45901089 | C=0.996, T=0.004 | C | C |
| rs13079478 | chr3:46007823 | G=0.995, T=0.005 | G | G |
| rs13059238 | chr3:46007825 | T=0.981, C=0.019 | T | C |
| rs33910087 | chr3:46009487 | G=0.983, A=0.017 | G | A |
| | | Haplotype Count | 1295 | 16 |
| | | Haplotype Frequency | 0.9796 | 0.0121 |

#### E. Asian

| RS Number | Position (GRCh37) | Allele Frequencies | Haplotypes |
|---|---|---|---|
| rs73064425 | chr3:45901089 | C=0.995, T=0.005 | C |
| rs13079478 | chr3:46007823 | G=0.996, T=0.004 | G |
| rs13059238 | chr3:46007825 | T=0.996, C=0.004 | T |
| rs33910087 | chr3:46009487 | G=0.995, A=0.005 | G |
| | | Haplotype Count | 1002 |
| | | Haplotype Frequency | 0.994 |

#### Amr

| RS Number | Position (GRCh37) | Allele Frequencies | Haplotypes | | |
|---|---|---|---|---|---|
| rs73064425 | chr3:45901089 | C=0.957, T=0.043 | C | T | C |
| rs13079478 | chr3:46007823 | G=0.941, T=0.059 | G | T | T |
| rs13059238 | chr3:46007825 | T=0.937, C=0.063 | T | C | C |
| rs33910087 | chr3:46009487 | G=0.941, A=0.059 | G | A | A |
| | | Haplotype Count | 644 | 24 | 17 |
| | | Haplotype Frequency | 0.928 | 0.0346 | 0.0245 |

#### CEU

| RS Number | Position (GRCh37) | Allele Frequencies | Haplotypes | | | |
|---|---|---|---|---|---|---|
| rs73064425 | chr3:45901089 | C=0.92, T=0.08 | C | T | C | T |
| rs13079478 | chr3:46007823 | G=0.878, T=0.122 | G | T | T | G |
| rs13059238 | chr3:46007825 | T=0.877, C=0.123 | T | C | C | T |
| rs33910087 | chr3:46009487 | G=0.878, A=0.122 | G | A | A | G |
| | | Haplotype Count | 871 | 69 | 54 | 11 |
| | | Haplotype Frequency | 0.8658 | 0.0686 | 0.0537 | 0.0109 |

#### S. Asian

| RS Number | Position (GRCh37) | Allele Frequencies | Haplotypes | | | |
|---|---|---|---|---|---|---|
| rs73064425 | chr3:45901089 | C=0.706, T=0.294 | C | T | C | T |
| rs13079478 | chr3:46007823 | G=0.64, T=0.36 | G | T | T | G |
| rs13059238 | chr3:46007825 | T=0.641, C=0.359 | T | C | C | T |
| rs33910087 | chr3:46009487 | G=0.641, A=0.359 | G | A | A | G |
| | | Haplotype Count | 561 | 222 | 128 | 65 |
| | | Haplotype Frequency | 0.5736 | 0.227 | 0.1309 | 0.0665 |
